# Drug–Drug Interaction Signals Between Anti-Amyloid-β Antibodies and Antithrombotic Drugs for Amyloid-Related Imaging Abnormalities and Intracranial Hemorrhage: A FAERS Disproportionality Analysis

**DOI:** 10.64898/2026.09.08.26362402

**Authors:** Tsuyoshi Nakai, Takenao Koseki, Hirohisa Watanabe, Shigeki Yamada

## Abstract

**Background:** Anti-amyloid-β (Aβ) monoclonal antibodies are disease-modifying treatments for early Alzheimer’s disease but are associated with amyloid-related imaging abnormalities (ARIA) and intracranial hemorrhage (ICH). However, the safety of concomitant antithrombotic use remains unclear.

**Objective:** To evaluate potential drug–drug interaction (DDI) signals between anti-Aβ antibodies and antithrombotic drugs for ARIA and ICH using the FDA Adverse Event Reporting System (FAERS; JAPIC AERS).

**Methods:** FAERS reports from 1997 to 2024 were analyzed. The anti-Aβ antibodies evaluated were aducanumab, lecanemab, and donanemab. Antithrombotics were aspirin, P2Y_12_ inhibitors, direct oral anticoagulants, warfarin, and tissue-type plasminogen activators (tPAs). Outcomes were any ARIA, ARIA with edema or effusion (ARIA-E), ARIA with hemosiderin deposition (ARIA-H), and ICH. Individual-drug signals were assessed using reporting odds ratios and information components; DDIs were evaluated using four complementary models.

**Results:** All three antibodies showed positive signals for all outcomes. When the three antibodies were analyzed together, aspirin showed positive DDI signals for any ARIA, ARIA-E, and ARIA-H in three models. Lecanemab–aspirin also showed signals for any ARIA and ARIA-H in multiple models. For ICH, tPAs showed signals in all four models with lecanemab and in the analysis combining all anti-Aβ antibodies. Both analyses were based on the same five lecanemab reports.

**Conclusions:** DDI signals were detected for anti-Aβ antibody–aspirin combinations for ARIA and anti-Aβ antibody–tPA combinations for ICH, particularly with lecanemab. These findings may warrant careful review of aspirin indications and baseline hemorrhagic risk and caution with thrombolysis during anti-Aβ antibody treatment, although prospective validation is needed.

## Introduction

Alzheimer’s disease (AD) is a progressive neurodegenerative disorder characterized by cognitive and functional decline, with cerebral amyloid-β (Aβ) deposition and tau pathology as its major neuropathological features.^1^ Pharmacological treatment of AD has traditionally relied on acetylcholinesterase (AChE) inhibitors, including donepezil, galantamine, and rivastigmine, and the *N*-methyl-*D*-aspartate (NMDA) receptor antagonist memantine. These drugs can improve cognition and daily functioning but do not target the underlying Aβ or tau pathology.^2^ Anti-Aβ monoclonal antibodies, including aducanumab, lecanemab, and donanemab, have recently been developed as disease-modifying treatments for early AD by targeting Aβ pathology in the brain. Phase 3 trials showed reductions in brain amyloid burden with all three antibodies and slower cognitive and functional decline with lecanemab and donanemab,^2–4^ whereas the two aducanumab trials yielded discordant clinical efficacy results.^5^

Despite their effects on brain amyloid burden, anti-Aβ antibodies are associated with amyloid-related imaging abnormalities (ARIA) and intracranial hemorrhage (ICH). ARIA is classified into ARIA with edema or effusion (ARIA-E) and ARIA with hemosiderin deposition (ARIA-H).^6–8^ ARIA-E is characterized by vasogenic edema or sulcal effusion, whereas ARIA-H includes cerebral microhemorrhages and cortical superficial siderosis. When symptomatic, ARIA may be serious or life-threatening.^7,9,10^ Although ICH was uncommon in clinical trials, fatal cases of ICH have been reported during treatment with lecanemab and donanemab.^9,10^ In clinical practice, anti-Aβ antibodies may be used concomitantly with antiplatelet or anticoagulant therapy in patients with cardiovascular or cerebrovascular comorbidities. Secondary analyses of clinical trials with lecanemab and donanemab reported broadly similar ARIA frequencies among participants with and without antithrombotic use.^9,10^ However, these analyses did not allow detailed assessment of individual antithrombotic drugs, as the small number of major hemorrhagic events limited the evaluation of drug-specific risks.^9,10^ Thrombolytic therapy may also be considered when acute ischemic stroke occurs during anti-Aβ antibody treatment. In one such case, multiple cerebral hemorrhages developed after intravenous tissue-type plasminogen activator (tPA) administration in a patient receiving lecanemab, resulting in death.^11^ Thus, the safety of specific anti-Aβ antibody–antithrombotic combinations requires further investigation.

The US Food and Drug Administration Adverse Event Reporting System (FAERS) is a large spontaneous-reporting database used for post-marketing safety surveillance, including exploratory signal detection and hypothesis generation.^12^ Recent FAERS-based studies have described adverse event (AE) reporting patterns for aducanumab, lecanemab, and donanemab, including signals for ARIA and hemorrhagic events.^13–16^ However, these studies mainly focused on individual antibodies and did not evaluate potential drug–drug interaction (DDI) signals between anti-Aβ antibodies and specific antithrombotic drugs and drug classes. Accordingly, this study aimed to evaluate potential DDI signals between the three anti-Aβ antibodies, including aducanumab, lecanemab, and donanemab, and major antithrombotic therapies for any ARIA, ARIA-E, ARIA-H, and ICH using FAERS data. Reporting signals for the study drugs were initially assessed individually, and potential DDIs were then evaluated between the anti-Aβ antibodies and aspirin, P2Y_12_ inhibitors, direct oral anticoagulants (DOACs), warfarin, and tPAs using four complementary signal-detection models.

## Methods

### Study design and data source

This pharmacovigilance study evaluated potential DDI signals between anti-amyloid-β antibodies and antithrombotic drugs for any ARIA, ARIA-E, ARIA-H, and ICH. Data were obtained from the Japan Pharmaceutical Information Center (JAPIC) AERS database (Tokyo, Japan), which is based on the FAERS database. The study was reported in accordance with the guidelines outlined in the REporting of A Disproportionality analysis for drUg Safety signal detection using individual case safety reports in PharmacoVigilance (READUS-PV) statement.^17^

### Dataset construction and preprocessing

The JAPIC AERS database contained the FAERS reports submitted from the fourth quarter of 1997 to the fourth quarter of 2024. The database had been preprocessed by JAPIC through the removal of duplicate case reports, unifying units, standardization of drug names using the WHO Drug Dictionary and Drug@FDA (https://www.accessdata.fda.gov), and assignment of the Medical Dictionary for Regulatory Activities/Japanese (MedDRA/J, https://www.jmo.gr.jp/jmo/servlet/mdrLoginTop) and Anatomical Therapeutic Chemical classifications. For this study, demographic, drug, and AE data were obtained from the DEMO, DRUG, and REAC tables, respectively. Cases with unknown sex or age data in the DEMO table were excluded. Duplicate data in the DRUG and REAC tables were removed. The DEMO, DRUG, and REAC tables were subsequently linked using PRIMARYID to create a relational database. In total, 11,258,022 cases were included in the disproportionality analyses (Figure 1). The JAPIC AERS database was processed using Navicat 17 in SQLite ver. 17.3.9 (PremiumSoft, Osaka, Japan).

**Figure 1.**
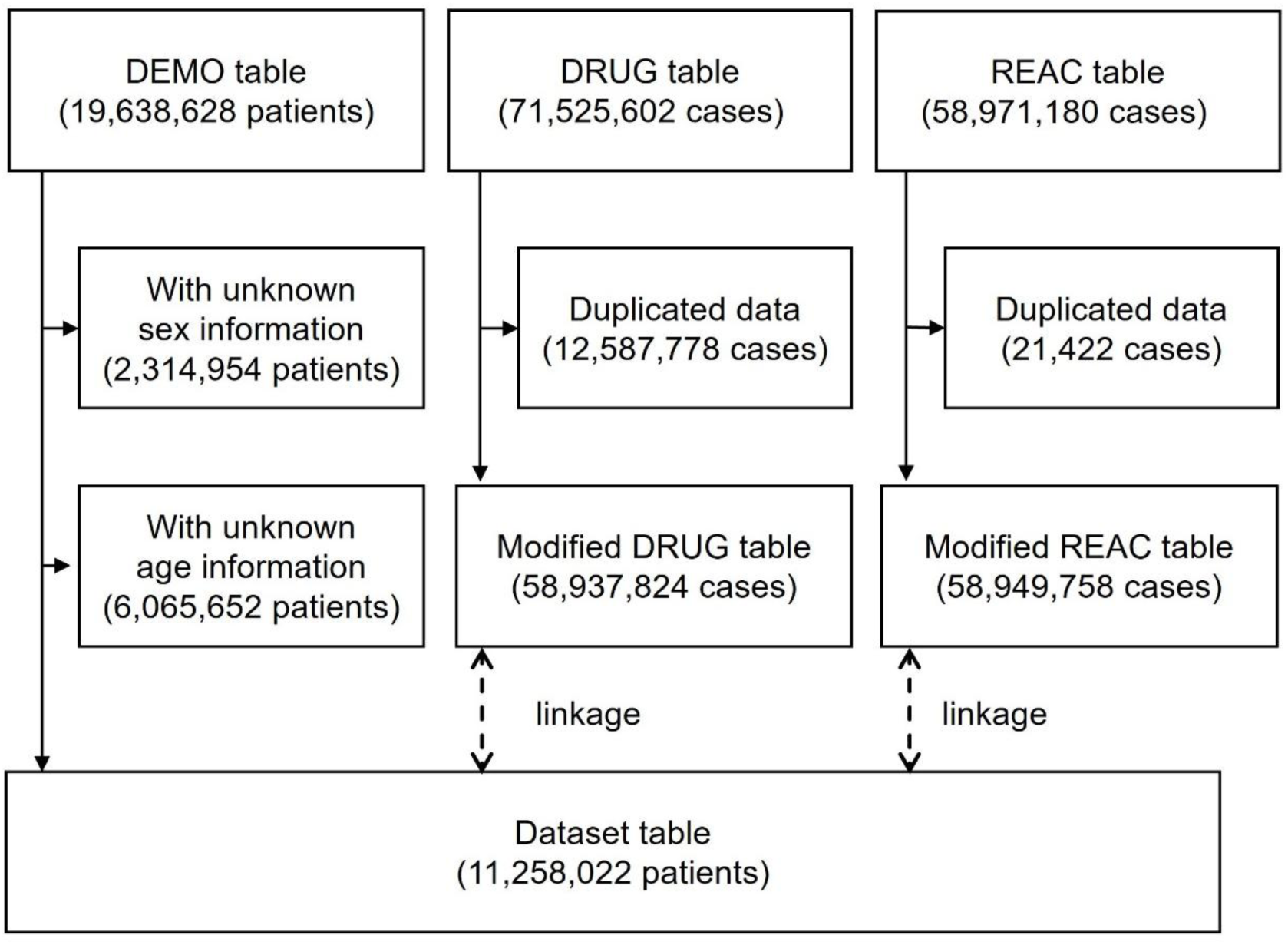
Flow diagram of dataset construction and preprocessing for the FDA Adverse Event Reporting System analysis.

### Target drugs

The anti-Aβ monoclonal antibodies evaluated were aducanumab, lecanemab, and donanemab. Other anti-dementia drugs were evaluated only in the monotherapy disproportionality analyses: the AChE inhibitors donepezil, galantamine, and rivastigmine and the NMDA receptor antagonist memantine. Antithrombotic drugs were classified into five groups: aspirin (acetylsalicylic acid), a cyclooxygenase-1 inhibitor; P2Y_12_ inhibitors, including clopidogrel, ticlopidine, prasugrel, and ticagrelor; DOACs, including apixaban, dabigatran, edoxaban, and rivaroxaban; warfarin, a vitamin K antagonist; and tPAs, including alteplase and tenecteplase. The substance IDs used to identify the study drugs in the JAPIC AERS database are listed in Supplemental Table 1. In the DRUG table, each drug record was classified by its reported role in relation to the reported AE as “primary suspect drug,” “secondary suspect drug,” “concomitant,” and “interacting.” Records from all four categories were included in the analyses.

### Definition of target adverse events

Any ARIA, ARIA-E, ARIA-H, and ICH were evaluated as target AEs. Relevant AEs were identified from the REAC table using preferred terms (PTs) coded according to the MedDRA/J (version 28.0). Any ARIA was defined as a composite outcome based on the presence of at least one of the four ARIA-related PTs listed in Supplemental Table 2. ARIA-E was defined by the presence of the PT listed in Supplemental Table 3, whereas ARIA-H was defined by the presence of at least one of the two PTs listed in Supplemental Table 4. ICH was defined by the presence of at least one PT included in the Haemorrhagic Central Nervous System Vascular Conditions Standardised MedDRA Query (SMQ; code 20000064; Supplemental Table 5). ARIA-H and ICH were evaluated separately because ARIA-H was identified using ARIA-specific PTs, whereas ICH was identified using the Haemorrhagic Central Nervous System Vascular Conditions SMQ.

### Signal detection and DDI analyses

To enhance the robustness of signal detection, complementary approaches were applied for individual-drug and DDI analyses. For the individual-drug analyses, reports involving concomitant use of two or more drugs evaluated in this study were excluded. AE reporting signals for any ARIA, ARIA-E, ARIA-H, and ICH were evaluated for monotherapy using the reporting odds ratio (ROR) and information component (IC).^18,19^ The RORs, ICs, and their 95% confidence intervals (CIs) were calculated using the two-by-two contingency table shown in Supplemental Table 6. An AE signal was considered positive when the lower limit of the 95% CI was > 1 for the ROR and > 0 for the IC.^20^

Potential DDI signals between anti-Aβ monoclonal antibodies and antithrombotic drugs were evaluated using four complementary methods: the Ω shrinkage measure model, additive model, multiplicative model, and chi-square statistics model.^20–22^ In each model, a positive DDI signal indicated that the reporting pattern for the drug combination exceeded that expected under the corresponding no-interaction assumption. These analyses were based on the four-by-two or two-by-two contingency tables shown in Supplemental Table 7. All calculations were performed using Microsoft Excel for Microsoft 365 (Microsoft Corporation, Redmond, WA, USA). The heatmap summarizing the number of models with positive DDI signals was generated using GraphPad Prism version 8.4.3 (GraphPad Software, San Diego, CA, USA).

### Reporting odds ratio

The ROR is a disproportionality measure that compares the reporting odds of a target AE for the target drug with those for all other drugs.^13–16,18^ The ROR and its 95% CI were calculated using the two-by-two contingency table shown in Supplemental Table 6 and Equations (1) and (2). An ROR signal was considered positive when the lower limit of the 95% CI exceeded 1.

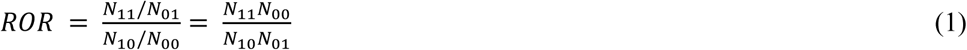

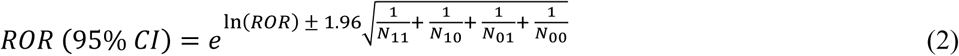

### Information components

The IC is a Bayesian disproportionality measure calculated using the Bayesian Confidence Propagation Neural Network method.^13–16,19^ This method incorporates statistical shrinkage, which can stabilize estimates when the number of reports is limited.^23^ The IC and its 95% CI were calculated using the two-by-two contingency table shown in Supplemental Table 6 and Equations (3)–(6). An IC signal was considered positive when the lower limit of the 95% CI exceeded 0.

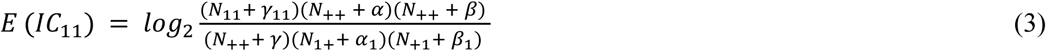

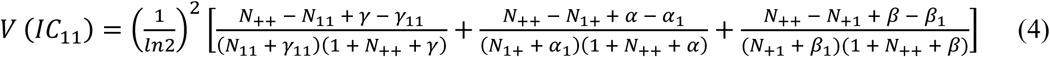

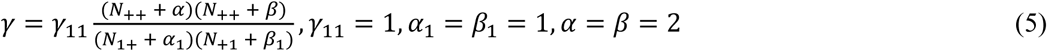

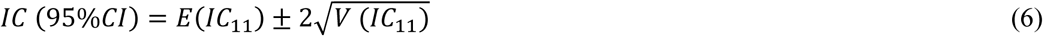

### Ω shrinkage measure model

The Ω shrinkage measure is a DDI signal detection method based on the ratio of the observed to the expected number of reports of a target AE under concomitant use of drugs *D*_1_ and *D*_2_.^24^ *Ω* and *Ω*_025_ were calculated using the four-by-two contingency table shown in Supplemental Table 7-1 and Equations (7) and (8).

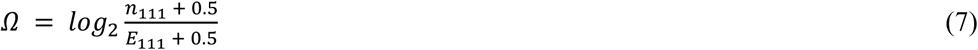

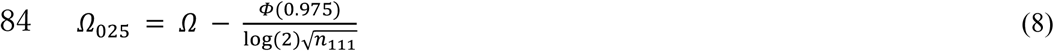

where *n*_111_ denotes the observed number of reports of the target AE under concomitant use of drugs *D*_1_ and *D*_2_, *E*_111_ denotes the corresponding expected number under the assumption of no interaction, and *Φ*(0.975) denotes the 97.5^th^ percentile of the standard normal distribution. A DDI signal was considered positive when *Ω*_025_ > 0.

### Additive model

Under the additive assumption, no interaction was assumed when the excess reporting proportion associated with concomitant use of drugs *D*_1_ and *D*_2_ was equal to the sum of the excess reporting proportions associated with each drug alone.^25^ The additive model was evaluated using the two-by-two contingency table shown in Supplemental Table 7-2 and Equation (9).

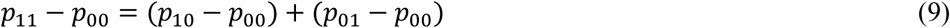

An additive DDI signal was considered positive when *p*_11_ − *p*_10_ − *p*_01_ + *p*_00_ > 0.

### Multiplicative model

Under the multiplicative model, no interaction was assumed when the relative reporting proportion associated with concomitant use of drugs *D*_1_and *D*_2_ was equal to the product of the relative reporting proportions associated with each drug alone.^25^ The multiplicative model was evaluated using the two-by-two contingency table shown in Supplemental Table 7-2 and Equation (10).

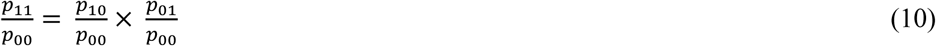

A multiplicative DDI signal was considered positive when 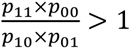.

### Chi-square statistics model

The chi-square statistics model compares the observed and expected numbers of reports of the target AE for the combination of drugs *D*_1_ and *D*_2_ under the assumption of no interaction.^26^ The χ was calculated using the four-by-two contingency table shown in Supplemental Table 7-1 and Equation (11).

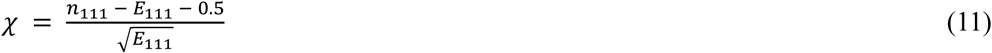

A DDI signal was considered positive when χ > 2.

## Results

### Population characteristics

The characteristics of the final analytical dataset are shown in Table 1. Of the 11,258,022 cases included in the dataset, 443 were classified as any ARIA cases and 11,257,579 as non-cases. Patients aged ≥ 65 years accounted for 87.1% of cases of any ARIA and 36.0% of non-cases. Female cases represented 58.9% of cases of any ARIA and 59.7% of non-cases. Anti-Aβ antibody use was reported in 414 cases of any ARIA (93.5%) and 1177 non-cases (0.01%).

**Table 1.** Characteristics of reported cases in the analysis of the FDA Adverse Event Reporting System.

|  | Total<br>(n=11,258,022) | Any ARIA |  |
| --- | --- | --- | --- |
|  |  | Cases<br>(n = 443) | Non-cases<br>(n = 11,257,579) |
| Age group, years, n (%) |  |  |  |
| < 65 | 7,200,404 (64.0) | 57 (12.9) | 7,200,347 (64.0) |
| ≥ 65 | 4,057,618 (36.0) | 386 (87.1) | 4,057,232 (36.0) |
| Sex, n (%) |  |  |  |
| Male | 4,541,494 (40.3) | 182 (41.1) | 4,541,312 (40.3) |
| Female | 6,716,528 (59.7) | 261 (58.9) | 6,716,267 (59.7) |
| Any anti-Aβ antibody reported, n (%) | 1,591 (0.01) | 414 (93.5) | 1,177 (0.01) |

Any ARIA was defined by the presence of at least one of the four preferred terms listed in Supplemental

### Reporting signals for individual drugs

The RORs and ICs for any ARIA, ARIA-E, ARIA-H, and ICH in the individual-drug analyses are summarized in Supplemental Tables 8–11. Among anti-Aβ antibodies, positive reporting signals were detected for aducanumab, lecanemab, and donanemab for all four target AEs. For any ARIA, positive signals were detected for aducanumab (ROR, 22,222.96 [95% CI 15,103.92 to 32,697.46]; IC, 5.64 [95% CI 5.13 to 6.15]), lecanemab (ROR, 8,766.00 [95% CI 7,143.95 to 10,756.33]; IC, 7.34 [95% CI 7.06 to 7.62]), and donanemab (ROR, 8,031.94 [95% CI 2,929.60 to 22,020.74]; IC, 2.58 [95% CI 1.25 to 3.92]). Positive reporting signals for ARIA-E and ARIA-H were also detected for all three anti-Aβ antibodies. In contrast, no positive reporting signals for any ARIA, ARIA-E, or ARIA-H were detected for AChE inhibitors, memantine, or any antithrombotic drug. For ICH, positive reporting signals were detected for all drugs evaluated in the individual-drug analyses.

### DDI signals between anti-Aβ antibodies and antithrombotic drugs

To reduce potential instability in signal estimates due to the limited number of reports for some individual antithrombotic drugs, antithrombotic drugs were grouped into five categories for the DDI analyses: aspirin, P2Y_12_ inhibitors, DOACs, warfarin, and tPAs. The DDI signal values between anti-Aβ antibodies and antithrombotic drugs for any ARIA, ARIA-E, ARIA-H, and ICH are summarized in Tables 2–5, and the number of models yielding positive DDI signals for each drug combination and target AE is summarized in Figure 2. For any ARIA, positive DDI signals were observed between all anti-Aβ antibodies combined and aspirin in the Ω shrinkage measure, additive, and chi-square statistics models (the signal value based on the Ω shrinkage measure model [Ω_025_] = 0.33; the signal value based on the additive model [AM] = 0.15; the signal value based on the chi-square statistics model [χ] = 3.75). Among the individual antibodies, lecanemab with aspirin showed positive signals in the Ω shrinkage measure and additive models (Ω_025_ = 0.01; AM = 0.10). Donanemab with DOACs also showed positive signals in the additive and multiplicative models (AM = 0.79; the signal value based on the multiplicative model [MM] = 5.85), although this finding was based on only one report involving this combination. No other combinations showed positive signals in multiple models.

**Figure 2.**
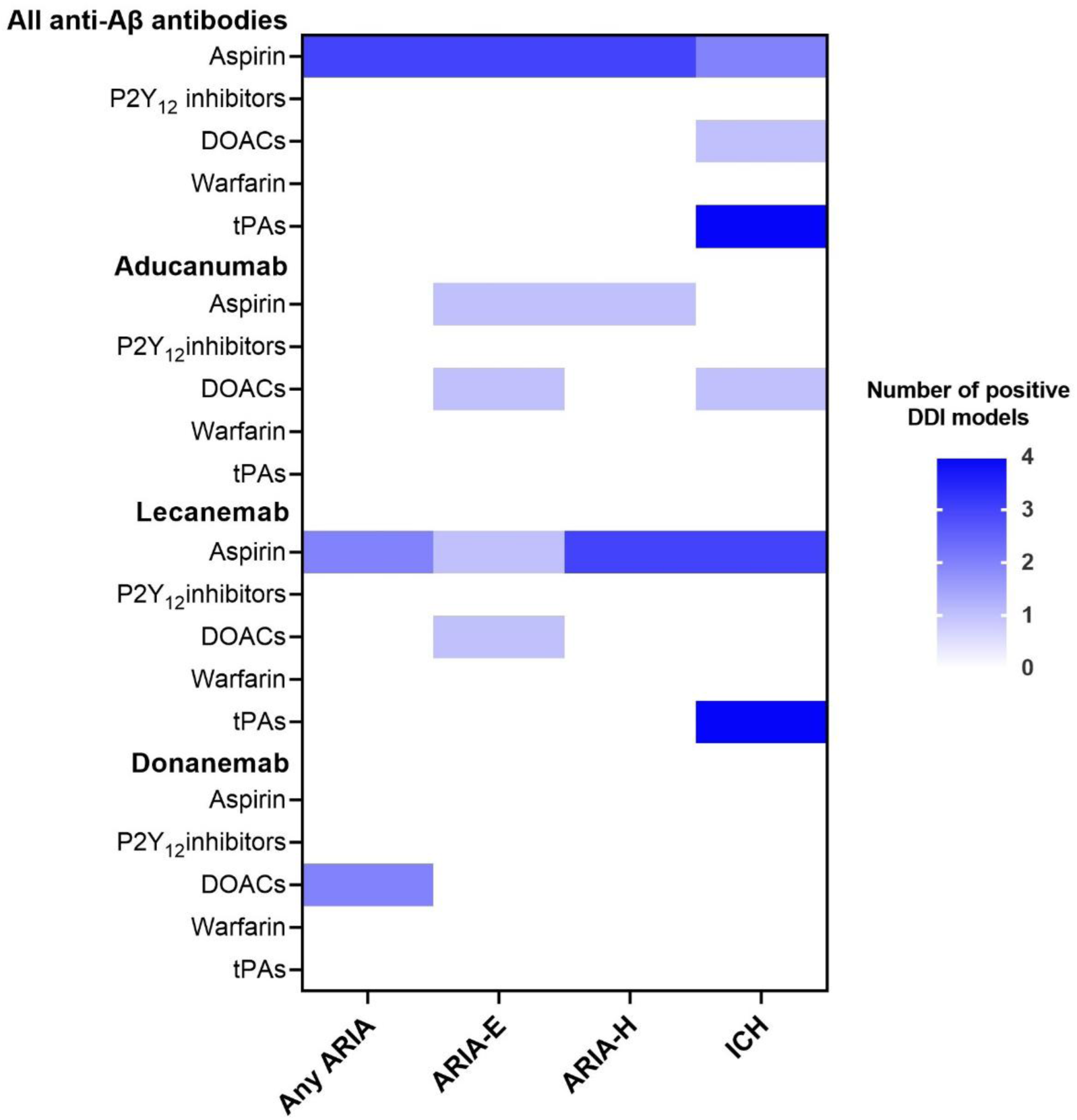
Heatmap summary of drug–drug interaction signals between anti-amyloid-β antibodies and antithrombotic drugs for any ARIA, ARIA-E, ARIA-H, and ICH. Each cell indicates the number of the four complementary DDI models that yielded a positive signal for each anti-Aβ antibody–antithrombotic drug combination and target AE (range, 0–4). Darker shading indicates a greater number of models yielding positive DDI signals. All anti-Aβ antibodies refers to aducanumab, lecanemab, and donanemab analyzed together. DDI: drug–drug interaction; AE: adverse event; DOACs: direct oral anticoagulants; tPAs: tissue-type plasminogen activators; ARIA: amyloid-related imaging abnormalities; ARIA-E: ARIA with edema or effusion; ARIA-H: ARIA with hemosiderin deposition; ICH: intracranial hemorrhage.

**Table 2.**
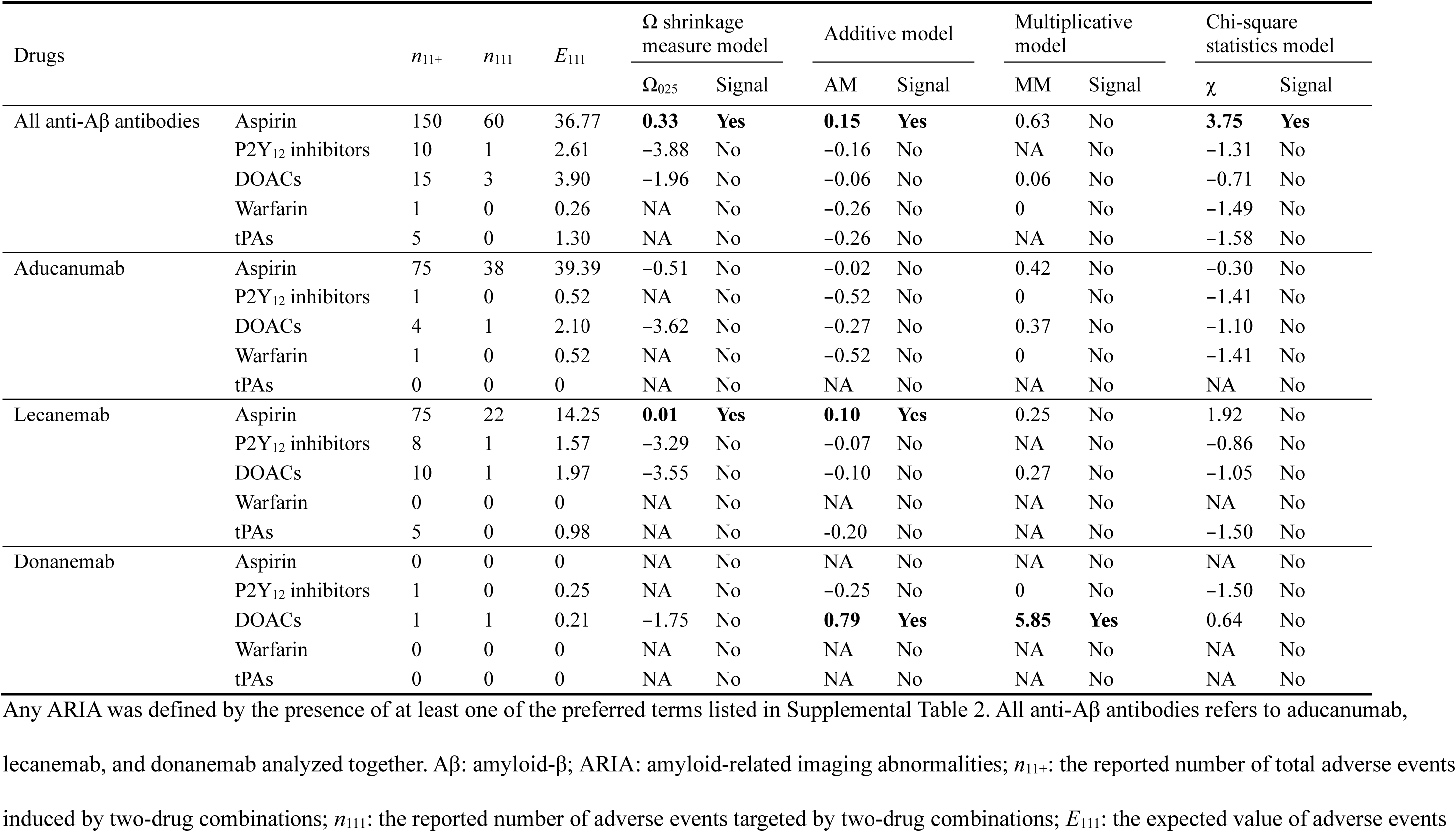
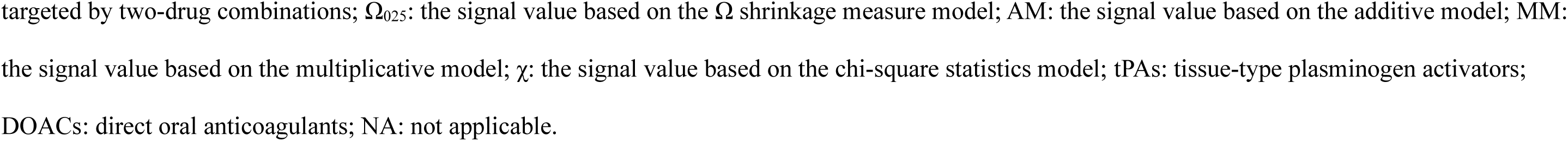
Drug–drug interaction signals of anti-Aβ antibodies with antithrombotic drugs for any ARIA.

**Table 3.**
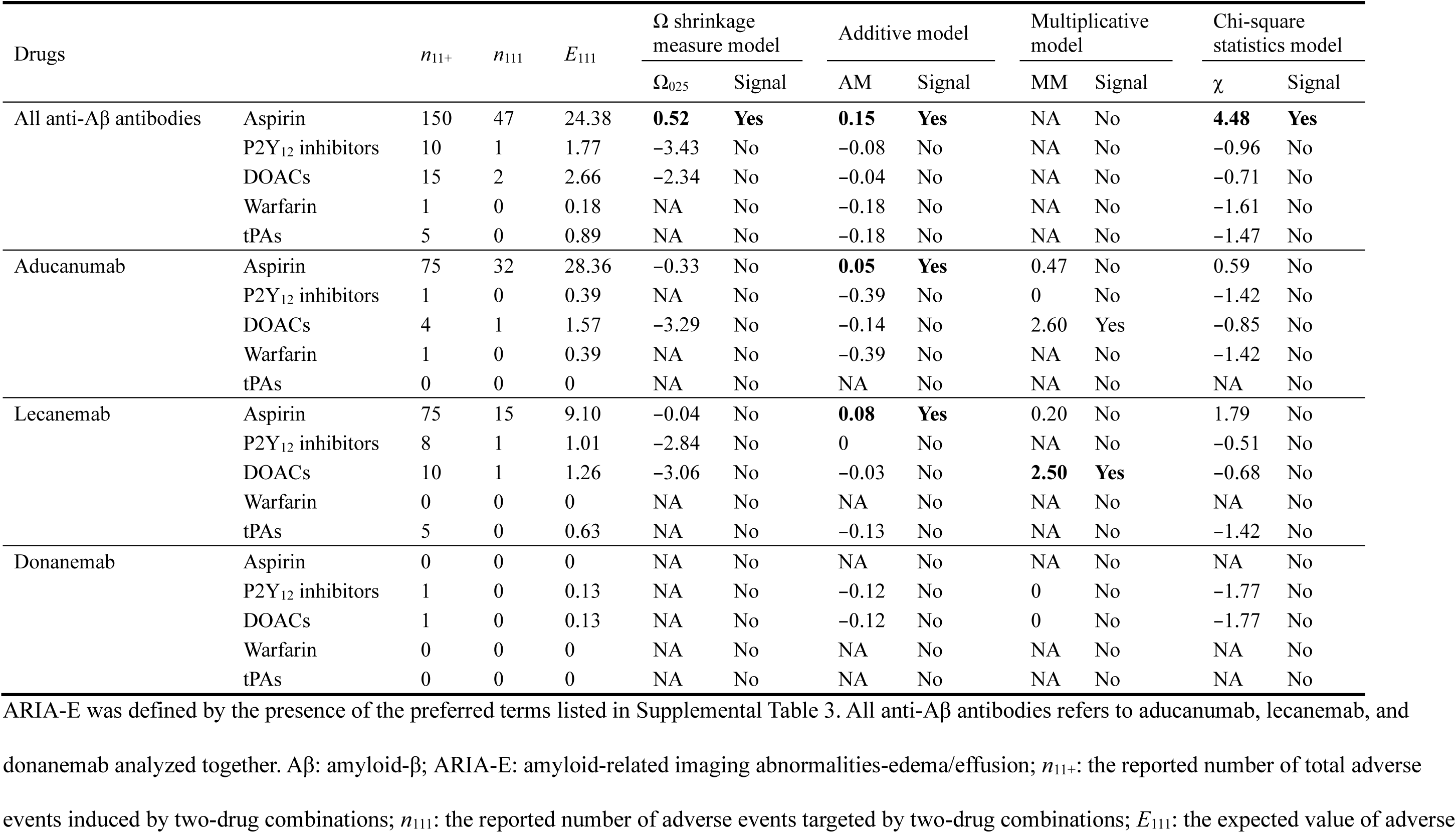
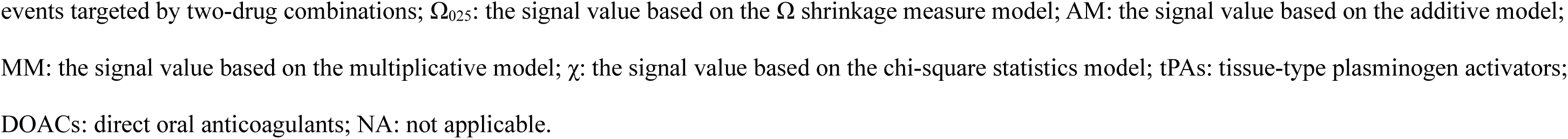
Drug–drug interaction signals of anti-Aβ antibodies with antithrombotic drugs for ARIA-E.

**Table 4.**
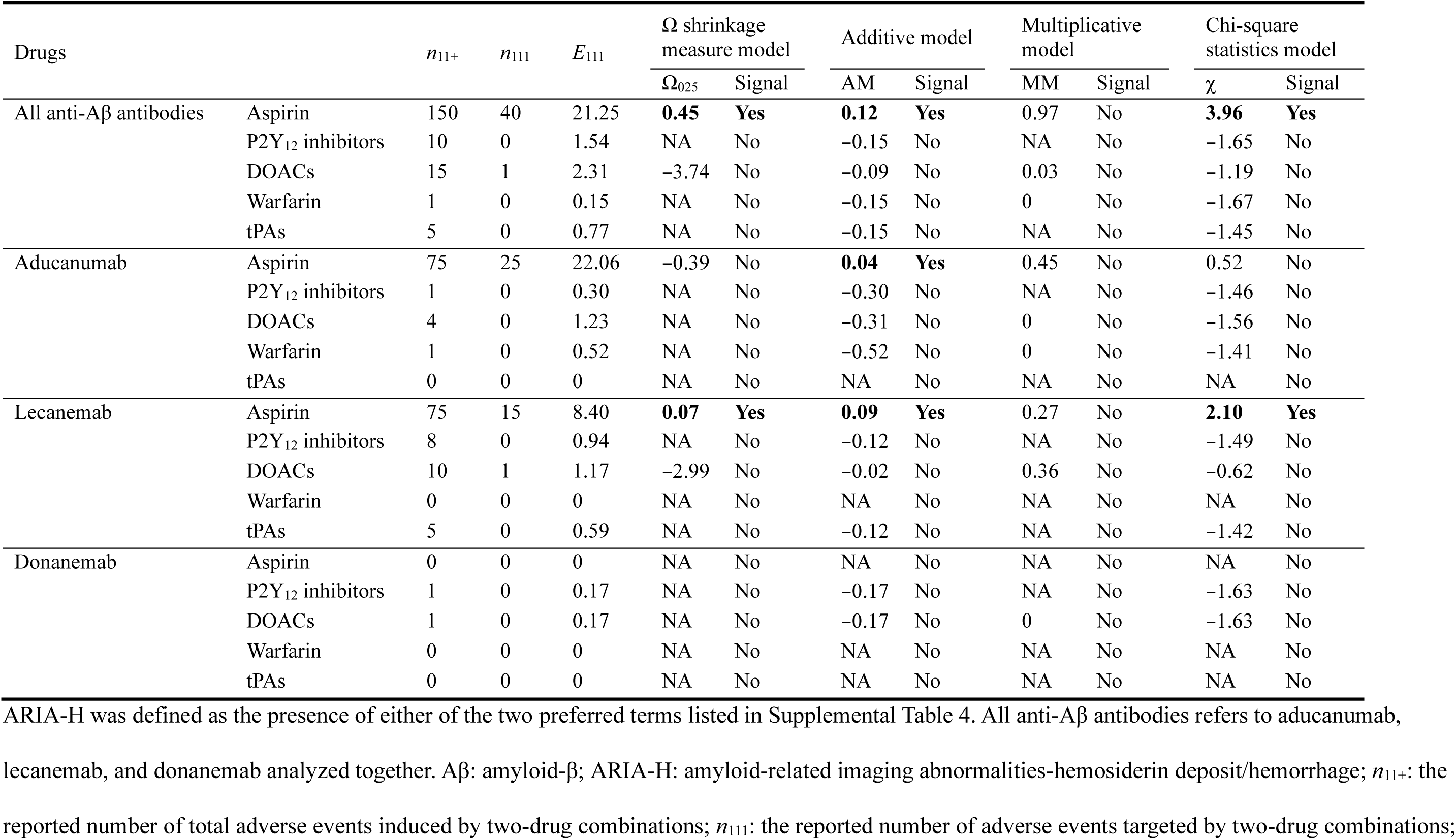
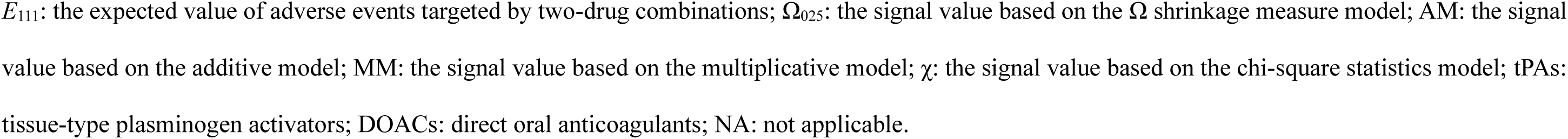
Drug–drug interaction signals of anti-Aβ antibodies with antithrombotic drugs for ARIA-H.

**Table 5.**
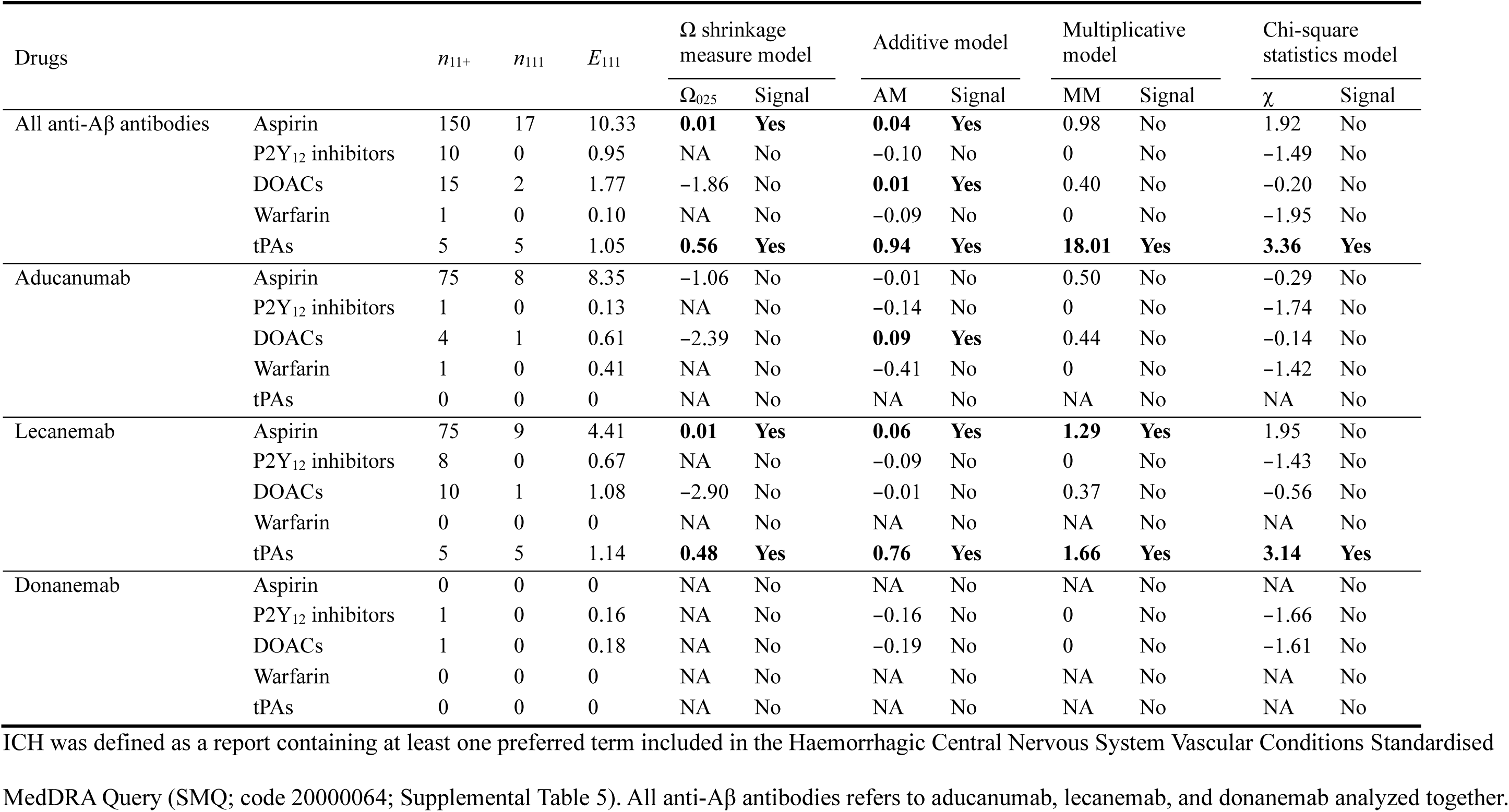
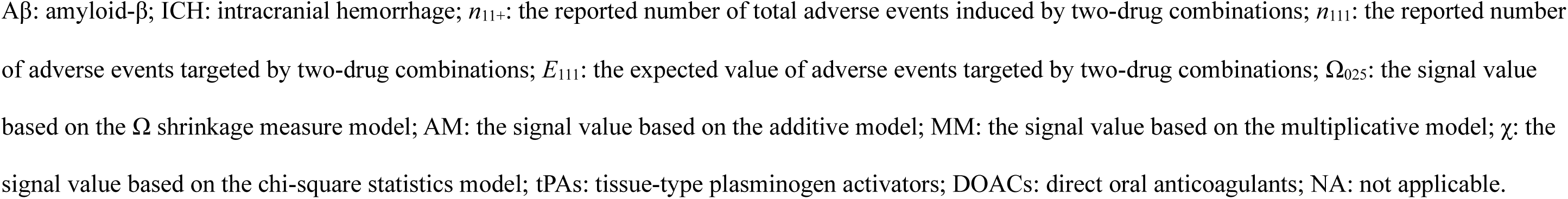
Drug–drug interaction signals of anti-Aβ antibodies with antithrombotic drugs for ICH.

For ARIA-E, all anti-Aβ antibodies combined showed positive DDI signals with aspirin in the Ω shrinkage measure, additive, and chi-square statistics models (Ω_025_ = 0.52; AM = 0.15; χ = 4.48). No individual antibody–antithrombotic drug combination showed positive signals in more than one model.

For ARIA-H, a similar pattern was observed for aspirin. All anti-Aβ antibodies combined showed positive DDI signals with aspirin in the Ω shrinkage measure, additive, and chi-square statistic models (Ω_025_ = 0.45; AM = 0.12; χ = 3.96). Additionally, lecanemab with aspirin showed positive signals in the same three models (Ω_025_ = 0.07; AM = 0.09; χ = 2.10). No other combinations showed positive signals in multiple models.

For ICH, the most consistent positive DDI signals were observed for combinations involving tPAs. Lecanemab with tPAs showed positive signals in all four models (Ω_025_ = 0.48; AM = 0.76; MM = 1.66; χ = 3.14). The same pattern was observed when all three anti-Aβ antibodies were analyzed together (Ω_025_ = 0.56; AM = 0.94; MM = 18.01; χ = 3.36); all five reports underlying these signals involved lecanemab. Positive DDI signals were also observed for lecanemab with aspirin in the Ω shrinkage measure, additive, and multiplicative models (Ω_025_ = 0.01; AM = 0.06; MM = 1.29), whereas aspirin showed positive signals only in the Ω shrinkage measure and additive models when all three anti-Aβ antibodies were analyzed together (Ω_025_ = 0.01; AM = 0.04). No combination with P2Y_12_ inhibitors, DOACs, or warfarin showed positive DDI signals in multiple models.

## Discussion

In this study, we evaluated potential DDI signals between anti-Aβ antibodies and major antithrombotic drug classes for ARIA and ICH using the FAERS database. The main findings were as follows. First, aducanumab, lecanemab, and donanemab showed positive reporting signals for any ARIA, ARIA-E, ARIA-H, and ICH in the individual-drug analyses. Second, aspirin showed positive DDI signals across ARIA outcomes when all three anti-Aβ antibodies were analyzed together. Among the antibody-specific analyses, the lecanemab–aspirin combination showed the most consistent signal pattern for any ARIA and ARIA-H. Third, tPAs showed the most consistent positive DDI signals for ICH, with all five reports underlying these signals involving lecanemab. Taken together, these DDI signals suggest greater-than-expected reporting under the corresponding no-interaction assumptions of multiple models, raising the possibility of combination-specific interactions.

In the individual-drug analyses, all three anti-Aβ antibodies showed positive reporting signals for any ARIA, ARIA-E, ARIA-H, and ICH, whereas no positive ARIA-related signals were detected for AChE inhibitors, memantine, or any of the antithrombotic drugs evaluated. These findings are consistent with the results of phase 3 clinical trials showing that ARIA occurred during treatment with aducanumab, lecanemab, and donanemab.^3–5^ Previous FAERS-based studies have similarly identified prominent ARIA-related signals for lecanemab,^13,16^ aducanumab,^14^ and donanemab.^15^ Unlike the ARIA-related outcomes, positive ICH signals were detected for all drugs included in the individual-drug analyses and were therefore not specific to anti-Aβ antibodies. Post-marketing analyses have also identified hemorrhage-related signals for lecanemab, aducanumab, and donanemab.^13–16^ These previous studies mainly characterized the post-marketing safety profiles of individual antibodies or compared PT-level reporting patterns across the three anti-Aβ antibodies. By contrast, this study evaluated ICH as a prespecified grouped outcome and examined potential DDI signals with specific antithrombotic drug classes. These individual-drug findings provide context for interpreting the aspirin- and tPA-related DDI signals described below.

In the DDI analyses, aspirin showed positive signals for any ARIA, ARIA-E, and ARIA-H when all anti-Aβ antibodies were analyzed together. Among the individual antibodies, the lecanemab–aspirin combination showed positive signals in multiple models for any ARIA and ARIA-H, whereas the ARIA-E signal was limited to the additive model. For aducanumab with aspirin, positive signals were confined to the additive model for ARIA-E and ARIA-H, and no aspirin reports were available for donanemab. Additionally, aspirin alone did not show an ARIA-related reporting signal. No comparable DDI signal pattern was observed with the other antithrombotic drug groups, except for the combination of donanemab with DOACs, which showed positive signals for any ARIA in two models. However, because this finding was based on a single report, it should be interpreted with caution. Secondary analyses of lecanemab and donanemab trials reported broadly similar ARIA frequencies among participants with and without concomitant antithrombotic use, and aspirin was the most commonly used antithrombotic drug.^9,10^ However, these analyses compared ARIA frequencies between participants with and without concomitant antithrombotic therapy and were not designed to assess potential interactions between specific drug combinations. Therefore, the present findings may provide complementary post-marketing information on potential DDI signals between aspirin and anti-Aβ antibodies across ARIA outcomes.

One possible explanation for the aspirin-related ARIA signals, particularly for ARIA-H, involves the combined effects of vascular vulnerability associated with Aβ clearance and aspirin-mediated platelet inhibition. Removal of vascular Aβ by anti-Aβ antibodies may transiently impair the integrity of amyloid-laden cerebral vessels and increase their susceptibility to vascular leakage or microhemorrhage, as suggested by experimental evidence and current pathophysiological models of ARIA.^7, 27^ In this setting, aspirin-mediated platelet inhibition may further impair hemostasis in vulnerable cerebral vessels. Although this mechanism more directly explains ARIA-H than ARIA-E, both often coexist and may share vascular injury and increased permeability as underlying pathological features. Therefore, the DDI signals observed across ARIA outcomes may reflect a broader influence of concomitant aspirin use on the vascular response to anti-Aβ antibody therapy. Although this mechanism is biologically plausible, FAERS data cannot determine whether concomitant aspirin use produces an additive or synergistic increase in ARIA risk. Accordingly, these findings should not be interpreted as supporting the routine withholding of aspirin when it is clinically indicated. Nevertheless, careful review of the indication for aspirin and the patient’s baseline hemorrhagic risk may be warranted before initiating anti-Aβ antibody therapy. Further studies using detailed clinical and imaging data are needed to clarify this association.

In the DDI analyses for ICH, aspirin showed positive ICH signals in multiple models, although less consistently than tPAs. These aspirin-related DDI signals may reflect the hemorrhagic vulnerability discussed above but should be interpreted with caution. A recent case report described a pontine hemorrhage that developed 20 days after the initiation of dual antiplatelet therapy with aspirin and clopidogrel in a patient who had received two lecanemab infusions.^28^ However, the specific contribution of aspirin could not be determined because clopidogrel was administered concomitantly. In the present analysis, no consistent DDI signal pattern was observed for P2Y_12_ inhibitors. Signals involving DOACs were confined to the additive model, and no consistent pattern was observed for warfarin.

The most consistent positive DDI signals for ICH involved tPAs. The lecanemab–tPA combination showed positive signals in all four models, and the same pattern was observed when all three anti-Aβ antibodies were analyzed together. However, all five reports contributing to these signals involved lecanemab; therefore, whether this finding extends to aducanumab or donanemab remains uncertain. A fatal case of multiple cerebral hemorrhages following intravenous tPA administration in a patient receiving lecanemab provides important clinical context for the present finding.^11^ tPAs activate plasminogen to plasmin and thereby promote fibrin degradation.^29^ In vessels affected by cerebral amyloid angiopathy or anti-Aβ antibody-associated vascular changes, this fibrinolytic effect may further impair local hemostasis and contribute to extensive ICH.^30^ The tPA-related DDI signals may also be clinically relevant to acute stroke care. Symptomatic ARIA can present with focal neurological deficits that mimic acute ischemic stroke, making rapid differentiation essential because decisions regarding reperfusion therapy are time-sensitive.^31–33^ Current appropriate use recommendations advise against thrombolytic treatment in patients receiving either lecanemab or donanemab.^31,32^ Additionally, the donanemab recommendations state that intravenous thrombolysis should not be administered without pretreatment magnetic resonance imaging (MRI) confirming the absence of ARIA.^32^ Accordingly, rapid confirmation of anti-Aβ antibody use and careful evaluation for ARIA are important when acute neurological symptoms occur during treatment.^31–33^ For eligible patients with large-vessel occlusion, mechanical thrombectomy without preceding thrombolysis may provide an alternative reperfusion strategy.^32,33^ However, the optimal management of patients who are not candidates for mechanical thrombectomy remains uncertain, and treatment decisions require individualized assessment of the potential neurological benefits and hemorrhagic risks.

This study has several limitations. First, FAERS does not provide denominator data and is susceptible to underreporting, reporting biases, and confounding. Reporting patterns may also have changed as ARIA became more widely recognized after anti-Aβ antibodies entered clinical practice. Thus, the findings from the disproportionality and DDI analyses cannot establish causality, quantify incidence or absolute risk, or demonstrate an additive or synergistic increase in clinical risk. They should not be used to directly compare clinical risks across drugs or drug combinations.^12,34^ Second, clinically relevant information on antithrombotic indications, doses, and the timing of drug exposure relative to event onset was incomplete or unavailable. Information on key ARIA risk factors, including APOE ε4 status and baseline MRI findings such as cerebral microhemorrhage burden, was also unavailable.^31,32^ Therefore, the effects of relevant clinical risk factors and the temporal relationship between drug exposure and event onset could not be adequately assessed. Third, ARIA and ICH were identified from MedDRA-coded reports and could not be independently confirmed using clinical records or neuroimaging. Therefore, misclassification, particularly between ARIA-H and other hemorrhagic events, cannot be excluded. Fourth, only a few reports were available for several antibody–antithrombotic combinations, and some combinations had no available reports. In particular, the ICH signals observed for combinations of anti-Aβ antibodies and tPAs were based on only five reports, all of which involved lecanemab, whereas data for combinations involving donanemab or warfarin were sparse. These limitations made it difficult to assess antibody-specific differences and whether the observed signals extend across the anti-Aβ antibody class. Although four complementary DDI models were used, differences in their underlying assumptions and sensitivity to sparse data may have contributed to discordant findings.^21,22^ Additionally, combining individual P2Y_12_ inhibitors, DOACs, and tPAs into drug classes may have obscured differences among individual drugs. Therefore, these findings should be interpreted as exploratory and hypothesis-generating. Further validation is needed in multicenter prospective cohort studies or large prospective registries that collect detailed data on antithrombotic exposure, standardized MRI findings, and relevant clinical risk factors, including APOE ε4 status.

## Conclusion

In this FAERS disproportionality analysis, potential DDI signals were identified between anti-Aβ antibodies and specific antithrombotic drugs. Aspirin showed potential DDI signals across ARIA-related outcomes, whereas tPAs showed the most consistent signals for ICH, particularly in combination with lecanemab. Across multiple models, these findings were consistent with greater-than-expected reporting under concomitant use and raised the possibility of combination-specific interactions. No similarly consistent DDI signal pattern was observed for P2Y_12_ inhibitors, DOACs, or warfarin, although the small number of reports limits comparisons across antithrombotic classes. Clinically, the aspirin-related findings may warrant careful review of the indication for aspirin and baseline hemorrhagic risk, whereas the tPA-related findings may support a cautious approach to thrombolytic therapy in patients receiving anti-Aβ antibodies. Because these findings are exploratory and do not establish causality or quantify absolute risk, further validation in prospective cohort studies or large treatment registries with detailed antithrombotic exposure data and standardized MRI assessments is needed.

## Data Availability

The FAERS datasets are publicly available from the FDA website. The datasets supporting the findings of this study can be obtained from the corresponding author upon reasonable request.

https://fis.fda.gov/extensions/FPD-QDE-FAERS/FPD-QDE-FAERS.html

## Acknowledgments

We want to thank Editage (www.editage.jp) for English language editing.

## Ethics considerations

Ethical approval was not required because this study used the open-access FAERS databases.

## Consent to participate

Consent to participate was not required because this study did not involve direct participation of human participants.

## Consent for publication

Not applicable.

## Author contribution(s)

**Tsuyoshi Nakai**: Conceptualization; Data curation; Formal analysis; Funding acquisition; Investigation; Methodology; Project administration; Validation; Visualization; Writing– original draft; Writing–review & editing.

**Takenao Koseki**: Data curation; Formal analysis; Investigation; Methodology; Writing– review & editing.

**Hirohisa Watanabe**: Formal analysis; Supervision; Writing– review & editing.

**Shigeki Yamada**: Formal analysis; Project administration; Supervision; Writing– review & editing.

## Funding

This work was supported by the Japan Society for the Promotion of Science (JSPS) KAKENHI (Grant Numbers: JP22K17824 and JP26K14596) for TN. The funder had no role in the study design, data collection and analysis, decision to publish, or preparation of the manuscript.

## Declaration of conflicting interests

The authors declared no potential conflicts of interest with respect to the research, authorship, and/or publication of this article.

## Data availability statement

The FAERS datasets are publicly available from the FDA website (https://fis.fda.gov/extensions/FPD-QDE-FAERS/FPD-QDE-FAERS.html). The datasets supporting the findings of this study can be obtained from the corresponding author upon reasonable request.

## Notes

### Competing Interest Statement

The authors have declared no competing interest.

### Author Declarations

JAPIC AERS is a preprocessed version of the publicly available FDA Adverse Event Reporting System (FAERS) database. Access to the preprocessed database was obtained through the Japan Pharmaceutical Information Center (JAPIC). The data were de-identified before the initiation of this study, and no identifiable patient information was accessed.

## References

1. Scheltens P, De Strooper B, Kivipelto M, et al. Alzheimer’s disease. Lancet 2021; 397: 1577–1590. DOI: 10.1016/s0140-6736(20)32205-4.

2. Fox NC, Belder C, Ballard C, et al. Treatment for Alzheimer’s disease. Lancet 2025; 406: 1408–1423. DOI: 10.1016/s0140-6736(25)01329-7.

3. van Dyck CH, Swanson CJ, Aisen P, et al. Lecanemab in early Alzheimer’s disease. N Engl J Med 2023; 388: 9–21. DOI: 10.1056/NEJMoa2212948.

4. Sims JR, Zimmer JA, Evans CD, et al. Donanemab in early symptomatic Alzheimer disease: The TRAILBLAZER-ALZ 2 Randomized Clinical Trial. Jama 2023; 330: 512–527. DOI: 10.1001/jama.2023.13239.

5. Budd Haeberlein S, Aisen PS, Barkhof F, et al. Two randomized phase 3 studies of aducanumab in early Alzheimer’s disease. J Prev Alzheimers Dis 2022; 9: 197–210. DOI: 10.14283/jpad.2022.30.

6. Sperling RA, Jack CR, Jr., Black SE, et al. Amyloid-related imaging abnormalities in amyloid-modifying therapeutic trials: recommendations from the Alzheimer’s Association Research Roundtable Workgroup. Alzheimers Dement 2011; 7: 367–385. DOI: 10.1016/j.jalz.2011.05.2351.

7. Hampel H, Elhage A, Cho M, et al. Amyloid-related imaging abnormalities (ARIA): radiological, biological and clinical characteristics. Brain 2023; 146: 4414–4424. DOI: 10.1093/brain/awad188.

8. Cogswell PM, Barakos JA, Barkhof F, et al. Amyloid-related imaging abnormalities with emerging Alzheimer disease therapeutics: Detection and reporting recommendations for clinical practice. AJNR Am J Neuroradiol 2022; 43: E19–E35. DOI: 10.3174/ajnr.A7586.

9. Zimmer JA, Ardayfio P, Wang H, et al. Amyloid-related imaging abnormalities with donanemab in early symptomatic Alzheimer disease: Secondary analysis of the TRAILBLAZER-ALZ and ALZ 2 randomized clinical trials. JAMA Neurol 2025; 82: 461–469. DOI: 10.1001/jamaneurol.2025.0065.

10. Honig LS, Sabbagh MN, van Dyck CH, et al. Updated safety results from phase 3 lecanemab study in early Alzheimer’s disease. Alzheimers Res Ther 2024; 16: 105. DOI: 10.1186/s13195-024-01441-8.

11. Reish NJ, Jamshidi P, Stamm B, et al. Multiple cerebral hemorrhages in a patient receiving lecanemab and treated with t-PA for stroke. N Engl J Med 2023; 388: 478–479. DOI: 10.1056/NEJMc2215148.

12. Potter E, Reyes M, Naples J, et al. FDA Adverse Event Reporting System (FAERS) Essentials: A guide to understanding, applying, and interpreting adverse event data reported to FAERS. Clin Pharmacol Ther 2025; 118: 567–582. DOI: 10.1002/cpt.3701.

13. Yan L, Zhang L, Xu Z, et al. A real-world disproportionality analysis of FDA adverse event reporting system (FAERS) events for lecanemab. Front Pharmacol 2025; 16: 1559447. DOI: 10.3389/fphar.2025.1559447.

14. Huang J, Long X, Chen C. A real-world safety surveillance study of aducanumab through the FDA adverse event reporting system. Front Pharmacol 2025; 16: 1522058. DOI: 10.3389/fphar.2025.1522058.

15. Zhang Y, Gong Q, Yu J, et al. Analysis signals of disproportionate reporting associated with donanemab: A retrospective pharmacovigilance study using the FAERS database. Sci Prog 2026; 109: 368504261461675. DOI: 10.1177/00368504261461675.

16. Xing X, Zhang X, Wang K, et al. Post-marketing safety concerns with lecanemab: a pharmacovigilance study based on the FDA Adverse Event Reporting System database. Alzheimers Res Ther 2025; 17: 15. DOI: 10.1186/s13195-024-01669-4.

17. Fusaroli M, Salvo F, Begaud B, et al. The Reporting of a Disproportionality Analysis for Drug Safety Signal Detection Using Individual Case Safety Reports in PharmacoVigilance (READUS-PV): Development and Statement. Drug Saf 2024; 47: 575–584. DOI: 10.1007/s40264-024-01421-9.

18. van Puijenbroek EP, Bate A, Leufkens HG, et al. A comparison of measures of disproportionality for signal detection in spontaneous reporting systems for adverse drug reactions. Pharmacoepidemiol Drug Saf 2002; 11: 3–10. DOI: 10.1002/pds.668.

19. Bate A, Lindquist M, Edwards IR, et al. A Bayesian neural network method for adverse drug reaction signal generation. Eur J Clin Pharmacol 1998; 54: 315–321. DOI: 10.1007/s002280050466.

20. Nakai T, Koseki T, Nakao H, et al. Analysis of hemorrhagic transformation and intracerebral hemorrhage under combination therapy with alteplase and antiplatelets or anticoagulants, using the Japanese Adverse Drug Event Report database. PLoS One 2025; 20: e0329378. DOI: 10.1371/journal.pone.0329378.

21. Noguchi Y, Tachi T, Teramachi H. Comparison of signal detection algorithms based on frequency statistical model for drug-drug interaction using spontaneous reporting systems. Pharm Res 2020; 37: 86. DOI: 10.1007/s11095-020-02801-3.

22. Noguchi Y, Tachi T, Teramachi H. Review of statistical methodologies for detecting drug-drug interactions using spontaneous reporting systems. Front Pharmacol 2019; 10: 1319. DOI: 10.3389/fphar.2019.01319.

23. Noguchi Y, Murayama A, Esaki H, et al. Angioedema caused by drugs that prevent the degradation of vasoactive peptides: A pharmacovigilance database study. J Clin Med 2021; 10. DOI: 10.3390/jcm10235507.

24. Norén GN, Sundberg R, Bate A, et al. A statistical methodology for drug-drug interaction surveillance. Stat Med 2008; 27: 3057–3070. DOI: 10.1002/sim.3247.

25. Thakrar BT, Grundschober SB, Doessegger L. Detecting signals of drug-drug interactions in a spontaneous reports database. Br J Clin Pharmacol 2007; 64: 489–495. DOI: 10.1111/j.1365-2125.2007.02900.x.

26. Gosho M, Maruo K, Tada K, et al. Utilization of chi-square statistics for screening adverse drug-drug interactions in spontaneous reporting systems. Eur J Clin Pharmacol 2017; 73: 779–786. DOI: 10.1007/s00228-017-2233-3.

27. Zago W, Schroeter S, Guido T, et al. Vascular alterations in PDAPP mice after anti-Aβ immunotherapy: Implications for amyloid-related imaging abnormalities. Alzheimers Dement 2013; 9: S105–S115. DOI: 10.1016/j.jalz.2012.11.010.

28. Chen S, Sun Y, Yao L, et al. Probable lecanemab-associated pontine hemorrhage following cardiovascular intervention: Clinical implications for lecanemab use. J Alzheimers Dis Rep 2025; 9: 25424823251366998. DOI: 10.1177/25424823251366998.

29. Collen D. Molecular mechanisms of fibrinolysis and their application to fibrin-specific thrombolytic therapy. J Cell Biochem 1987; 33: 77–86. DOI: 10.1002/jcb.240330202.

30. McCarron MO, Nicoll JA. Cerebral amyloid angiopathy and thrombolysis-related intracerebral haemorrhage. Lancet Neurol 2004; 3: 484–492. DOI: 10.1016/s1474-4422(04)00825-7.

31. Cummings J, Apostolova L, Rabinovici GD, et al. Lecanemab: Appropriate use recommendations. J Prev Alzheimers Dis 2023; 10: 362–377. DOI: 10.14283/jpad.2023.30.

32. Rabinovici GD, Selkoe DJ, Schindler SE, et al. Donanemab: Appropriate use recommendations. J Prev Alzheimers Dis 2025; 12: 100150. DOI: 10.1016/j.tjpad.2025.100150.

33. Greenberg SM, Bax F, van Veluw SJ. Amyloid-related imaging abnormalities: manifestations, metrics and mechanisms. Nat Rev Neurol 2025; 21: 193–203. DOI: 10.1038/s41582-024-01053-8.

34. Michel C, Scosyrev E, Petrin M, et al. Can disproportionality analysis of post-marketing case reports be used for comparison of drug safety profiles? Clin Drug Investig 2017; 37: 415–422. DOI: 10.1007/s40261-017-0503-6.

