## Supplemental Material for "Drug–Drug Interaction Signals Between Anti-Amyloid-β Antibodies and Antithrombotic Drugs for Amyloid-Related Imaging Abnormalities and Intracranial Hemorrhage: A FAERS Disproportionality Analysis"

Tsuyoshi Nakai, Ph.D.

ORCID: 0009-0005-2667-7057

Department of Pharmacotherapeutics and Informatics, Fujita Health University School of Medicine, 1-98 Dengakugakubo, Kutsukake-cho, Toyoake, Aichi 470-1192, Japan

**Supplemental material**

**Supplemental Table 1.** Study drugs and corresponding substance IDs**.**

| Pharmacological classification | Drugs | Substance IDs |
|---|---|---|
| **Anti-dementia drugs** | | |
| Anti-Aβ antibodies | Aducanumab | f0000001836, j0000400840 |
| | Lecanemab | f0000001899, j0000424005 |
| | Donanemab | f0000001966, j0000435160 |
| AChE inhibitors | Donepezil | w0000010218, w0000010219 |
| | Galantamine | w0000001743, w0000003224 |
| | Rivastigmine | j0000420563, w0000010283, w0000012092 |
| NMDA receptor antagonist | Memantine | w0000006204, w0000011218 |
| **Antiplatelets** | | |
| COX-1 inhibitor | Aspirin | w0000000051, w0000001518 |
| $P2Y_{12}$ inhibitors | Clopidogrel | f0000001180, j0000391302, j0000396859, j0000398058 |
| | Ticlopidine | w0000007763, w0000007900 |
| | Prasugrel | f0000001376, j0000423982, j0000430996, w0000016748 |
| | Ticagrelor | j0000365693 |
| **Anticoagulants** | | |
| DOACs | Apixaban | w0000018092 |
| | Rivaroxaban | w0000010779 |
| | Edoxaban | j0000399638, w0000018617 |
| | Dabigatran | j0000430882, w0000010779, w0000013175 |
| Vitamin K antagonist | Warfarin | w0000000702, w0000001299, w0000003414, w0000012848 |
| **Antithrombotic agents** | | |
| tPAs | Alteplase | w0000009955 |
| | Tenecteplase | w0000010738 |

Aβ: amyloid-β; AChE: acetylcholinesterase; NMDA: *N*-methyl-*D*-aspartate receptor; COX-1: cyclooxygenase-1; DOACs: direct oral anticoagulants; tPAs: tissue-type plasminogen activators.

**Supplemental Table 2.** Definition of any amyloid-related imaging abnormalities (ARIA).

| PT code | PT name |
|---|---|
| 10072599 | Amyloid related imaging abnormalities |
| 10072260 | Amyloid related imaging abnormality-oedema/effusion |
| 10072601 | Amyloid related imaging abnormality-microhaemorrhages and haemosiderin deposits |
| 10070564 | Superficial siderosis of central nervous system |

PT: preferred term.

**Supplemental Table 3.** Definition of amyloid-related imaging abnormalities-edema/effusion (ARIA-E).

| PT code | PT name |
|---|---|
| 10072260 | Amyloid related imaging abnormality-oedema/effusion |

PT: preferred term.

**Supplemental Table 4.** Definition of amyloid-related imaging abnormalities-hemosiderin deposit/hemorrhage (ARIA-H).

| PT code | PT name |
|---|---|
| 10072601 | Amyloid related imaging abnormality-microhaemorrhages and haemosiderin deposits |
| 10070564 | Superficial siderosis of central nervous system |

PT: preferred term.

**Supplemental Table 5.** Definition of intracranial hemorrhage (ICH).

| SMQ code | SMQ name |
|---|---|
| 20000064 | Haemorrhagic central nervous system vascular conditions (SMQ) |
| PT code | PT name |
| 10077031 | Basal ganglia haematoma |
| 10067057 | Basal ganglia haemorrhage |
| 10071043 | Basal ganglia stroke |
| 10075736 | Basilar artery perforation |
| 10073230 | Brain stem haematoma |
| 10006145 | Brain stem haemorrhage |
| 10071205 | Brain stem microhaemorrhage |
| 10068644 | Brain stem stroke |
| 10051328 | Carotid aneurysm rupture |
| 10075728 | Carotid artery perforation |
| 10072043 | Central nervous system haemorrhage |
| 10061038 | Cerebellar haematoma |
| 10008030 | Cerebellar haemorrhage |
| 10071206 | Cerebellar microhaemorrhage |
| 10079062 | Cerebellar stroke |
| 10075394 | Cerebral aneurysm perforation |
| 10008076 | Cerebral aneurysm ruptured syphilitic |
| 10008086 | Cerebral arteriovenous malformation haemorrhagic |
| 10075734 | Cerebral artery perforation |
| 10082099 | Cerebral cyst haemorrhage |
| 10053942 | Cerebral haematoma |
| 10008111 | Cerebral haemorrhage |
| 10050157 | Cerebral haemorrhage foetal |
| 10008112 | Cerebral haemorrhage neonatal |
| 10067277 | Cerebral microhaemorrhage |
| 10008190 | Cerebrovascular accident |
| 10008196 | Cerebrovascular disorder |
| 10073681 | Epidural haemorrhage |
| 10078254 | Extra-axial haemorrhage |
| 10015769 | Extradural haematoma |
| 10082797 | Extradural haematoma evacuation |

**Supplemental Table 5 (continued).**

| | |
|---|---|
| 10080347 | Extraischaemic cerebral haematoma |
| 10082594 | Foville syndrome |
| 10018985 | Haemorrhage intracranial |
| 10085944 | Haemorrhagic cerebellar infarction |
| 10019005 | Haemorrhagic cerebral infarction |
| 10019016 | Haemorrhagic stroke |
| 10055677 | Haemorrhagic transformation stroke |
| 10062025 | Intracerebral haematoma evacuation |
| 10059491 | Intracranial haematoma |
| 10086946 | Intracranial haemorrhage neonatal |
| 10022775 | Intracranial tumour haemorrhage |
| 10022840 | Intraventricular haemorrhage |
| 10022841 | Intraventricular haemorrhage neonatal |
| 10052593 | Meningorrhagia |
| 10027580 | Middle cerebral artery stroke |
| 10089110 | Occipital lobe stroke |
| 10089109 | Parietal lobe stroke |
| 10073945 | Perinatal stroke |
| 10076706 | Periventricular haemorrhage neonatal |
| 10056447 | Pituitary apoplexy |
| 10049760 | Pituitary haemorrhage |
| 10058940 | Putamen haemorrhage |
| 10039330 | Ruptured cerebral aneurysm |
| 10076051 | Spinal cord haematoma |
| 10048992 | Spinal cord haemorrhage |
| 10050162 | Spinal epidural haematoma |
| 10049236 | Spinal epidural haemorrhage |
| 10082031 | Spinal stroke |
| 10073564 | Spinal subarachnoid haemorrhage |
| 10050164 | Spinal subdural haematoma |
| 10073563 | Spinal subdural haemorrhage |
| 10090938 | Spontaneous subdural haematoma |
| 10059613 | Stroke in evolution |
| 10076701 | Subarachnoid haematoma |

**Supplemental Table 5 (continued).**

| | |
|---|---|
| 10042316 | Subarachnoid haemorrhage |
| 10042317 | Subarachnoid haemorrhage neonatal |
| 10089974 | Subcortical stroke |
| 10042361 | Subdural haematoma |
| 10042363 | Subdural haematoma evacuation |
| 10042364 | Subdural haemorrhage |
| 10042365 | Subdural haemorrhage neonatal |
| 10090985 | Thalamic microhaemorrhage |
| 10087626 | Thalamic stroke |
| 10058939 | Thalamus haemorrhage |
| 10075735 | Vertebral artery perforation |

PT: preferred term; SMQ: Standardised MedDRA Query.

**Supplemental Table 6.** Two-by-two contingency table for adverse-event signal detection.

| | Target AE | All other AEs | Total |
|---|---|---|---|
| Target drug | $N_{11}$ | $N_{10}$ | $N_{1+}$ |
| All other drugs | $N_{01}$ | $N_{00}$ | $N_{0+}$ |
| Total | $N_{+1}$ | $N_{+0}$ | $N_{++}$ |

AEs: adverse events.

**Supplemental Table 7.** Contingency tables for drug–drug interaction signal detection.

**7-1.** Four-by-two contingency table for drug–drug interaction signal detection.

| | Target AE | All other AEs | Total |
|---|---|---|---|
| Target drug $D_1$ and $D_2$ | $n_{111}$ | $n_{110}$ | $n_{11+}$ |
| Only drug $D_1$ | $n_{101}$ | $n_{100}$ | $n_{10+}$ |
| Only drug $D_2$ | $n_{011}$ | $n_{010}$ | $n_{01+}$ |
| Neither drug $D_1$ nor drug $D_2$ | $n_{001}$ | $n_{000}$ | $n_{00+}$ |
| Total | $n_{++1}$ | $n_{++0}$ | $n_{+++}$ |

AEs: adverse events.

**7-2.** Two-by-two contingency table for drug–drug interaction signal detection.

| | With drug $D_2$ | Without drug $D_2$ |
|---|---|---|
| With drug $D_1$ | $p_{11} = n_{111}/n_{11+}$ | $p_{10} = n_{101}/n_{10+}$ |
| Without drug $D_1$ | $p_{01} = n_{011}/n_{01+}$ | $p_{00} = n_{001}/n_{00+}$ |

**Supplemental Table 8.** Reporting odds ratio and information components of any ARIA for each drug as monotherapy.

| Pharmacological classification | Drugs | $N_{11}$ | $N_{10}$ | $N_{01}$ | $N_{00}$ | ROR [95% CI] | IC [95% CI] | Signal |
|---|---|---|---|---|---|---|---|---|
| **Anti-dementia drugs** | | | | | | | | |
| Anti-Aβ antibodies | Aducanumab | 49 | 63 | 394 | 11,257,516 | **22,222.96 [15,103.92 to 32,697.46]** | **5.64 [5.13 to 6.15]** | **Yes** |
| | Lecanemab | 167 | 777 | 276 | 11,256,802 | **8,766.00 [7,143.95 to 10,756.33]** | **7.34 [7.06 to 7.62]** | **Yes** |
| | Donanemab | 5 | 16 | 438 | 11,257,563 | **8,031.94 [2,929.60 to 22,020.74]** | **2.58 [1.25 to 3.92]** | **Yes** |
| AChE inhibitors | Donepezil | 0 | 15,743 | 443 | 11,241,836 | 0 | −0.70 [−3.59 to 2.19] | No |
| | Galantamine | 0 | 2,757 | 443 | 11,254,822 | 0 | −0.15 [−3.04 to 2.74] | No |
| | Rivastigmine | 0 | 10,245 | 443 | 11,247,334 | 0 | −0.49 [−3.38 to 2.40] | No |
| NMDA receptor antagonist | Memantine | 0 | 8,488 | 443 | 11,249,091 | 0 | −0.42 [−3.31 to 2.47] | No |
| **Antiplatelets** | | | | | | | | |
| COX-1 inhibitor | Aspirin | 3 | 452,862 | 440 | 10,804,717 | 0.16 [0.05 to 0.51] | −2.24 [−3.69 to −0.79] | No |
| $P2Y_{12}$ inhibitors | Clopidogrel | 0 | 42,353 | 443 | 11,215,226 | 0 | −1.42 [−4.31 to 1.47] | No |
| | Ticlopidine | 0 | 4,636 | 443 | 11,252,943 | 0 | −0.24 [−3.13 to 2.65] | No |
| | Prasugrel | 0 | 3,083 | 443 | 11,254,496 | 0 | −0.17 [−3.05 to 2.72] | No |
| | Ticagrelor | 0 | 8,452 | 443 | 11,249,127 | 0 | −0.42 [−3.30 to 2.47] | No |
| **Anticoagulants** | | | | | | | | |
| DOACs | Apixaban | 1 | 132,212 | 442 | 11,125,367 | 0.19 [0.03 to 1.35] | −1.64 [−3.68 to 0.41] | No |
| | Rivaroxaban | 5 | 95,163 | 438 | 11,162,416 | 1.34 [0.55 to 3.23] | 0.34 [−0.85 to 1.52] | No |
| | Edoxaban | 0 | 7,624 | 443 | 11,249,955 | 0 | −0.38 [−3.27 to 2.51] | No |
| | Dabigatran | 1 | 43,218 | 442 | 11,214,361 | 0.59 [0.08 to 4.18] | −0.44 [−2.48 to 1.61] | No |
| Vitamin K antagonist | Warfarin | 2 | 144,226 | 441 | 11,113,353 | 0.35 [0.09 to 1.40] | −1.16 [−2.83 to 0.51] | No |
| **Antithrombotic agents** | | | | | | | | |
| tPAs | Alteplase | 0 | 9,258 | 443 | 11,248,321 | 0 | −0.45 [−3.34 to 2.44] | No |
| | Tenecteplase | 0 | 771 | 443 | 11,256,808 | 0 | −0.04 [−2.93 to 2.85] | No |

**Supplemental Table 8 (continued).**

$N_{11}$, $N_{10}$, $N_{01}$, and $N_{00}$ are shown in Supplemental Table 6, respectively. Any ARIA was defined by the presence of at least one of the preferred terms listed in Supplemental Table 2. ARIA: amyloid-related imaging abnormalities; ROR: reporting odds ratio; IC: information components; CI: confidence interval; Aβ: amyloid-β; AChE: acetylcholinesterase; NMDA: *N*-methyl-*D*-aspartate receptor; COX-1: cyclooxygenase-1; DOACs: direct oral anticoagulants; tPAs: tissue-type plasminogen activators.

**Supplemental Table 9.** Reporting odds ratio and information components of ARIA-E for each drug as monotherapy.

| Pharmacological classification | Drugs | $N_{11}$ | $N_{10}$ | $N_{01}$ | $N_{00}$ | ROR [95% CI] | IC [95% CI] | Signal |
|---|---|---|---|---|---|---|---|---|
| **Anti-dementia drugs** | | | | | | | | |
| Anti-Aβ antibodies | Aducanumab | 31 | 81 | 251 | 11,257,659 | **17,165.29 [11,143.46 to 26,441.25]** | **5.00 [4.39 to 5.60]** | **Yes** |
| | Lecanemab | 105 | 839 | 177 | 11,256,901 | **7,959.26 [6,194.11 to 10,227.45]** | **6.69 [6.35 to 7.04]** | **Yes** |
| | Donanemab | 3 | 18 | 279 | 11,257,722 | **6,725.04 [1,969.82 to 22,959.53]** | **2.00 [0.42 to 3.58]** | **Yes** |
| AChE inhibitors | Donepezil | 0 | 15,743 | 282 | 11,241,997 | 0 | −0.48 [−3.37 to 2.41] | No |
| | Galantamine | 0 | 2,757 | 282 | 11,254,983 | 0 | −0.10 [−2.99 to 2.79] | No |
| | Rivastigmine | 0 | 10,245 | 282 | 11,247,495 | 0 | −0.33 [−3.22 to 2.56] | No |
| NMDA receptor antagonist | Memantine | 0 | 8,488 | 282 | 11,249,252 | 0 | −0.28 [−3.17 to 2.61] | No |
| **Antiplatelets** | | | | | | | | |
| COX-1 inhibitor | Aspirin | 0 | 452,865 | 282 | 10,804,875 | 0 | −3.63 [−6.52 to −0.74] | No |
| $P2Y_{12}$ inhibitors | Clopidogrel | 0 | 42,353 | 282 | 11,215,387 | 0 | −1.05 [−3.94 to 1.84] | No |
| | Ticlopidine | 0 | 4,636 | 282 | 11,253,104 | 0 | −0.16 [−3.05 to 2.73] | No |
| | Prasugrel | 0 | 3,083 | 282 | 11,254,657 | 0 | −0.11 [−3.00 to 2.78] | No |
| | Ticagrelor | 0 | 8,452 | 282 | 11,249,288 | 0 | −0.28 [−3.17 to 2.61] | No |
| **Anticoagulants** | | | | | | | | |
| DOACs | Apixaban | 0 | 132,213 | 282 | 11,125,527 | 0 | −2.11 [−5.00 to 0.78] | No |
| | Rivaroxaban | 0 | 95,168 | 282 | 11,162,572 | 0 | −1.76 [−4.65 to 1.13] | No |
| | Edoxaban | 0 | 7,624 | 282 | 11,250,116 | 0 | −0.25 [−3.14 to 2.64] | No |
| | Dabigatran | 0 | 43,219 | 282 | 11,214,521 | 0 | −1.06 [−3.95 to 1.83] | No |
| Vitamin K antagonist | Warfarin | 0 | 144,228 | 282 | 11,113,512 | 0 | −2.21 [−5.10 to 0.68] | No |
| **Antithrombotic agents** | | | | | | | | |
| tPAs | Alteplase | 0 | 9,258 | 282 | 11,248,482 | 0 | −0.30 [−3.19 to 2.59] | No |
| | Tenecteplase | 0 | 771 | 282 | 11,256,969 | 0 | −0.03 [−2.92 to 2.86] | No |

**Supplemental Table 9 (continued).**

$N_{11}$, $N_{10}$, $N_{01}$, and $N_{00}$ are shown in Supplemental Table 6, respectively. ARIA-E was defined by the presence of the preferred terms listed in Supplemental Table 3. ARIA-E: amyloid-related imaging abnormalities-edema/effusion; ROR: reporting odds ratio; IC: information components; CI: confidence interval; Aβ: amyloid-β; AChE: acetylcholinesterase; NMDA: *N*-methyl-*D*-aspartate receptor; COX-1: cyclooxygenase-1; DOACs: direct oral anticoagulants; tPAs: tissue-type plasminogen activators.

**Supplemental Table 10.** Reporting odds ratio and information components of ARIA-H for each drug as monotherapy.

| Pharmacological classification | Drugs | $N_{11}$ | $N_{10}$ | $N_{01}$ | $N_{00}$ | ROR [95% CI] | IC [95% CI] | Signal |
|---|---|---|---|---|---|---|---|---|
| **Anti-dementia drugs** | | | | | | | | |
| Anti-Aβ antibodies | Aducanumab | 25 | 87 | 245 | 11,257,665 | **13,203.92 [8,318.26 to 20,959.15]** | **4.70 [4.04 to 5.35]** | **Yes** |
| | Lecanemab | 97 | 847 | 173 | 11,256,905 | **7,451.80 [5,759.66 to 9,641.07]** | **6.58 [6.23 to 6.94]** | **Yes** |
| | Donanemab | 4 | 17 | 266 | 11,257,735 | **9,958.19 [3,328.68 to 29,791.24]** | **2.32 [0.88 to 3.76]** | **Yes** |
| AChE inhibitors | Donepezil | 0 | 10,245 | 270 | 11,247,507 | 0 | −0.32 [−3.21 to 2.57] | No |
| | Galantamine | 0 | 2,757 | 270 | 11,254,995 | 0 | −0.09 [−2.98 to 2.80] | No |
| | Rivastigmine | 0 | 10,245 | 270 | 11,247,507 | 0 | −0.32 [−3.21 to 2.57] | No |
| NMDA receptor antagonist | Memantine | 0 | 8,488 | 270 | 11,249,264 | 0 | −0.27 [−3.16 to 2.62] | No |
| **Antiplatelets** | | | | | | | | |
| COX-1 inhibitor | Aspirin | 2 | 452,863 | 268 | 10,804,889 | 0.18 [0.04 to 0.72] | −1.99 [−3.66 to −0.31] | No |
| $P2Y_{12}$ inhibitors | Clopidogrel | 0 | 42,353 | 270 | 11,215,399 | 0 | −1.01 [−3.90 to 1.88] | No |
| | Ticlopidine | 0 | 4,636 | 270 | 11,253,116 | 0 | −0.15 [−3.04 to 2.74] | No |
| | Prasugrel | 0 | 3,083 | 270 | 11,254,669 | 0 | −0.10 [−2.99 to 2.79] | No |
| | Ticagrelor | 0 | 8,452 | 270 | 11,249,300 | 0 | −0.27 [−3.16 to 2.62] | No |
| **Anticoagulants** | | | | | | | | |
| DOACs | Apixaban | 1 | 132,212 | 269 | 11,125,540 | 0.31 [0.04 to 2.23] | −1.06 [−3.11 to 0.98] | No |
| | Rivaroxaban | 5 | 95,163 | 265 | 11,162,589 | 2.21 [0.91 to 5.36] | 0.87 [−0.32 to 2.06] | No |
| | Edoxaban | 0 | 7,624 | 270 | 11,250,128 | 0 | −0.24 [−3.13 to 2.65] | No |
| | Dabigatran | 1 | 43,218 | 269 | 11,214,534 | 0.96 [0.14 to 6.87] | −0.03 [−2.08 to 2.02] | No |
| Vitamin K antagonist | Warfarin | 2 | 144,226 | 268 | 11,113,526 | 0.58 [0.14 to 2.31] | −0.58 [−2.25 to 1.10] | No |
| **Antithrombotic agents** | | | | | | | | |
| tPAs | Alteplase | 0 | 9,258 | 270 | 11,248,494 | 0 | −0.29 [−3.18 to 2.60] | No |
| | Tenecteplase | 0 | 771 | 270 | 11,256,981 | 0 | −0.03 [−2.92 to 2.87] | No |

**Supplemental Table 10 (continued).**

$N_{11}$, $N_{10}$, $N_{01}$, and $N_{00}$ are shown in Supplemental Table 6, respectively. ARIA-H was defined as the presence of either of the two preferred terms listed in Supplemental Table 4. ARIA-H: amyloid-related imaging abnormalities-hemosiderin deposit/hemorrhage; ROR: reporting odds ratio; IC: information components; CI: confidence interval; Aβ: amyloid-β; AChE: acetylcholinesterase; NMDA: *N*-methyl-*D*-aspartate receptor; COX-1: cyclooxygenase-1; DOACs: direct oral anticoagulants; tPAs: tissue-type plasminogen activators.

**Supplemental Table 11.** Reporting odds ratio and information components of ICH for each drug as monotherapy.

| Pharmacological classification | Drugs | $N_{11}$ | $N_{10}$ | $N_{01}$ | $N_{00}$ | ROR [95% CI] | IC [95% CI] | Signal |
|---|---|---|---|---|---|---|---|---|
| **Anti-dementia drugs** | | | | | | | | |
| Anti-Aβ antibodies | Aducanumab | 9 | 103 | 171,897 | 11,086,013 | **5.64 [2.85 to 11.14]** | **1.88 [0.92 to 2.83]** | **Yes** |
| | Lecanemab | 38 | 906 | 171,868 | 11,085,210 | **2.71 [1.96 to 3.74]** | **1.34 [0.87 to 1.81]** | **Yes** |
| | Donanemab | 3 | 18 | 171,903 | 11,086,098 | **10.75 [3.17 to 36.49]** | **1.58 [0.01 to 3.15]** | **Yes** |
| AChE inhibitors | Donepezil | 384 | 15,359 | 171,522 | 11,070,757 | **1.61 [1.46 to 1.79]** | **0.67 [0.52 to 0.82]** | **Yes** |
| | Galantamine | 97 | 2,660 | 171,809 | 11,083,456 | **2.35 [1.92 to 2.88]** | **1.18 [0.89 to 1.48]** | **Yes** |
| | Rivastigmine | 420 | 9,825 | 171,486 | 11,076,291 | **2.76 [2.50 to 3.04]** | **1.42 [1.28 to 1.56]** | **Yes** |
| NMDA receptor antagonist | Memantine | 161 | 8,327 | 171,745 | 11,077,789 | **1.25 [1.07 to 1.46]** | **0.31 [0.08 to 0.54]** | **Yes** |
| **Antiplatelets** | | | | | | | | |
| COX-1 inhibitor | Aspirin | 14,441 | 438,424 | 157,465 | 10,647,692 | **2.23 [2.19 to 2.27]** | **1.06 [1.04 to 1.09]** | **Yes** |
| $P2Y_{12}$ inhibitors | Clopidogrel | 2,482 | 39,871 | 169,424 | 11,046,245 | **4.06 [3.90 to 4.23]** | **1.94 [1.88 to 2.00]** | **Yes** |
| | Ticlopidine | 188 | 4,448 | 171,718 | 11,081,668 | **2.73 [2.36 to 3.16]** | **1.40 [1.18 to 1.61]** | **Yes** |
| | Prasugrel | 194 | 2,889 | 171,712 | 11,083,227 | **4.33 [3.75 to 5.01]** | **2.02 [1.81 to 2.23]** | **Yes** |
| | Ticagrelor | 340 | 8,112 | 171,566 | 11,078,004 | **2.71 [2.43 to 3.02]** | **1.39 [1.23 to 1.55]** | **Yes** |
| **Anticoagulants** | | | | | | | | |
| DOACs | Apixaban | 8,421 | 123,792 | 163,485 | 10,962,324 | **4.56 [4.46 to 4.67]** | **2.06 [2.03 to 2.09]** | **Yes** |
| | Rivaroxaban | 8,132 | 87,036 | 163,774 | 10,999,080 | **6.27 [6.13 to 6.42]** | **2.48 [2.45 to 2.52]** | **Yes** |
| | Edoxaban | 363 | 7,261 | 171,543 | 11,078,855 | **3.23 [2.91 to 3.59]** | **1.63 [1.48 to 1.79]** | **Yes** |
| | Dabigatran | 5,002 | 38,217 | 166,904 | 11,047,899 | **8.66 [8.41 to 8.93]** | **2.92 [2.88 to 2.96]** | **Yes** |
| Vitamin K antagonist | Warfarin | 7,343 | 136,885 | 164,563 | 10,949,231 | **3.57 [3.48 to 3.66]** | **1.74 [1.70 to 1.77]** | **Yes** |
| **Antithrombotic agents** | | | | | | | | |
| tPAs | Alteplase | 1,880 | 7,378 | 170,026 | 11,078,738 | **16.60 [15.78 to 17.47]** | **3.72 [3.65 to 3.80]** | **Yes** |
| | Tenecteplase | 226 | 545 | 171,680 | 11,085,571 | **26.78 [22.93 to 31.27]** | **4.15 [3.93 to 4.37]** | **Yes** |

**Supplemental Table 11 (continued).**

$N_{11}$, $N_{10}$, $N_{01}$, and $N_{00}$ are shown in Supplemental Table 6, respectively. ICH was defined as a report containing at least one preferred term included in the Haemorrhagic Central Nervous System Vascular Conditions SMQ (Supplemental Table 5). ICH: intracranial hemorrhage; ROR: reporting odds ratio; IC: information components; CI: confidence interval; Aβ: amyloid-β; AChE: acetylcholinesterase; NMDA: *N*-methyl-*D*-aspartate receptor; COX-1: cyclooxygenase-1; DOACs: direct oral anticoagulants; tPAs: tissue-type plasminogen activators.
